# Health Circuit: a practice-proven adaptive case management approach for innovative healthcare services

**DOI:** 10.1101/2023.03.22.23287569

**Authors:** Carmen Herranz, Laura Martín, Fernando Dana, Antoni Sisó-Almirall, Josep Roca, Isaac Cano

## Abstract

Digital health tools may facilitate care continuum. However, enhancement of digital aid is imperative to prevent information gaps or redundancies, as well as to facilitate support of flexible care plans. The study presents Health Circuit, a digital health tool with an adaptive case management approach and analyses its healthcare impact, as well as its usability (SUS) and acceptability (NPS) by healthcare professionals and patients. In 2018-19, an initial prototype of Health Circuit was tested in a cluster randomized clinical pilot (n=100) in patients with high risk for hospitalization (Study I). In 2021, a pilot version of Health Circuit was evaluated in 104 high risk patients undergoing prehabilitation before major surgery (Study II). In study I, Health Circuit resulted in reduction of emergency room visits [4 (13%) vs 7 (44%)] and enhanced patients’ empowerment (p<0.0001) and showed good acceptability/usability scores (NPS 31 and SUS 54/100). In Study II, NPS scored 40 and SUS 85/100. The acceptance rate was also high (mean score of 8.4/10). Health Circuit showed potential for healthcare value generation, good both acceptability and usability despite being a prototype system, prompting the need for testing a completed system in real-world scenarios.

## 1. Introduction

Worldwide, conventional disease-oriented approaches are being replaced by innovative preventive patient-centered healthcare services with digital health tools supporting vertical (between the hospital and the community) and horizontal (between health and social services) integration. However, despite the broad consensus there are still significant challenges in ensuring care continuum across health organizations. ^1^

In many cases, cooperation among healthcare tiers and providers needs digital support for secure bi-directional and instantaneous communications to avoid information gaps or duplications and assist the care team in making shared decisions. Moreover, the non-deterministic nature of healthcare services requires a shift in the focus from a set of pre-scriptive tasks to a context-dependent, flexible, often evolving, care plans aimed at achieving optimal outcomes for each patient.

Back in 2001, Adaptive Case Management (ACM) was introduced in the workflow management literature to support knowledge workers (e.g., physicians, architects, lawyers, etc.) ^2^, as well as unpredictable business processes that require flexibility. Because knowledge-based work is data-intensive, the ACM approach can foster more informed decisions and customise business processes as necessary ^34^. The application of these ACM principles is essential for efficient digital health transformation. Improved end-user engagement, personalization to the evolving patient’s health status, and better communication and coordination between health care professionals are just some examples of the benefits of ACM when applied to innovative health services ^5^

The current study presents the pre-commercial experience of co-design and prototyping of a digital health tool with an ACM approach, Health Circuit, during the period 2018-2021. The process was composed of two consecutive steps. In an initial phase, 2018-2019, the feasibility of a secure bi-directional communication channel was tested for improving the management of complex chronic patients with high risk of hospitalization. In a second phase, ACM principles were incorporated (2019-2021) to enable virtual sharing of care plans among all team members of an innovative Prehabilitation service ^6–8^ including both home and community-based care, addressed to candidates for major abdominal surgeries.

While the first pilot study aimed to analyse its potential with respect to management of unplanned events and patient self-management, both studies shared as common objective the assessment of the ACM approach of Health Circuit with respect to its usability and acceptability as perceived by end users.

## 2. Materials and Methods

### 2.1 Context

It is of note that the pre-commercial experience reported in the current study benefited from the combined input of all the stakeholders, including end users, from a health district integrated care area in the city of Barcelona (AISBE; 520,000 citizens) ^9^ with a long-lasting experience in piloting digital support for enhanced management of chronic patients ^10,11^, falling within the activities of the Catalan Open Innovation Hub on Digitally Enabled Integrated Care Services, which is one of the four original EU Good Practices in the European Joint Action JADECARE ^12^

Within this regional digital health transformation context, the first reported study for prevention of hospitalizations in complex chronic patients (2019-2020) focused on assessment of a prototype-level (Technology Readiness Level - TRL 5) ^13^ secure bi-directional communication channel with a twofold purpose: (i) Facilitating management of unplanned events through a direct access to one case manager who could eventually trigger a shared decision making process with other health professionals (primary care, specialists, or both); and (ii) Empowering patients’ self-management aiming at increasing self-efficacy through shared videos or educational material.

The second reported study (2020-2021) focused on assessing the acceptability and usability of a pilot version of the Health Circuit approach (TRL 7) for enhancing patient adherence to the prehabilitation program at Hospital Clínic of Barcelona (HCB) ^14,15^. In recent years, prehabilitation has emerged as a health-valued intervention to reduce postoperative morbi-mortality in a variety of surgical populations ^16^. In this line, we have previously demonstrated efficacy ^7^ and potential effectiveness ^8,17^ of prehabilitation. It is of note that the impact of prehabilitation in real-world settings seems strongly associated to actionable areas such as enhancing patients’ adherence and transferability to community settings, which requires digital support with an ACM approach ^15^ to enable virtual sharing of care plans among professionals across health tiers and prescription of non-pharmacological interventions for patient empowerment for self-management.

### 2.2 Health Circuit approach

The ACM approach of Health Circuit aims to empower professionals to define dynamic service workflows as patient-centred evidence-based interventions with special input on combining pharmacological and non-pharmacological interventions. It can be viewed as a semi-automated and event-driven task manager for professionals and patients that enable them to perform certain activities in the context of the patients’ treatment.

It is collaborative in the sense that healthcare professionals must share their knowledge and decisions with their peers, which is achieved with the following main features (Error! Reference source not found.):

-Case-centered. The collaboration is bounded by the case, meaning that a group of professionals interact with each other and the patient in the context of the case.

-Multi-channel communication channel. The communication channel is a tool for helping in decision making, that is, a secure chat between N peers starts with a particular issue in mind, and, when resolved, leaves a summary of the decision and motivations for it, which then is translated into some action. It is also used to help patients solve doubts and issues with their treatment. This is the front-line support to collaborative work.

-Multi-disciplinary shared care plan. The activities to be performed are related to different specialties/disciplines from various levels of care but are shared in a common care plan. A care plan represents the workflow of how a treatment is planned to be addressed. It has a layered structure with three levels of elements, from bottom to top:

- Tasks. What is ultimately actionable. They represent things like “the 3rd session of physiotherapy at the hospital gym”, “filling by 2nd time some questionnaires”, “the 5th day of receiving nutritional tips”.
- Activities. Generate identical tasks over a period with a given recurrence. They can be started or ended automatically by linking them to other activities’ events.
- Groups of activities. Help grouping activities to be enabled or disabled at a higher level, e.g., baseline assessment, weekly follow-up, etc.
- The structural dependencies between the above elements, in the sense of the care plan or service workflow that represents the treatment, translate into two main concepts:
- Semi-automated processes. Some activities are dependent or related to the outcome and/or completion of other activities.
- Overridable. The structure does not impose strict rules to professionals, so they can override any decision with proper motivation, which is registered into the system for accountability.

**Figure 1.**
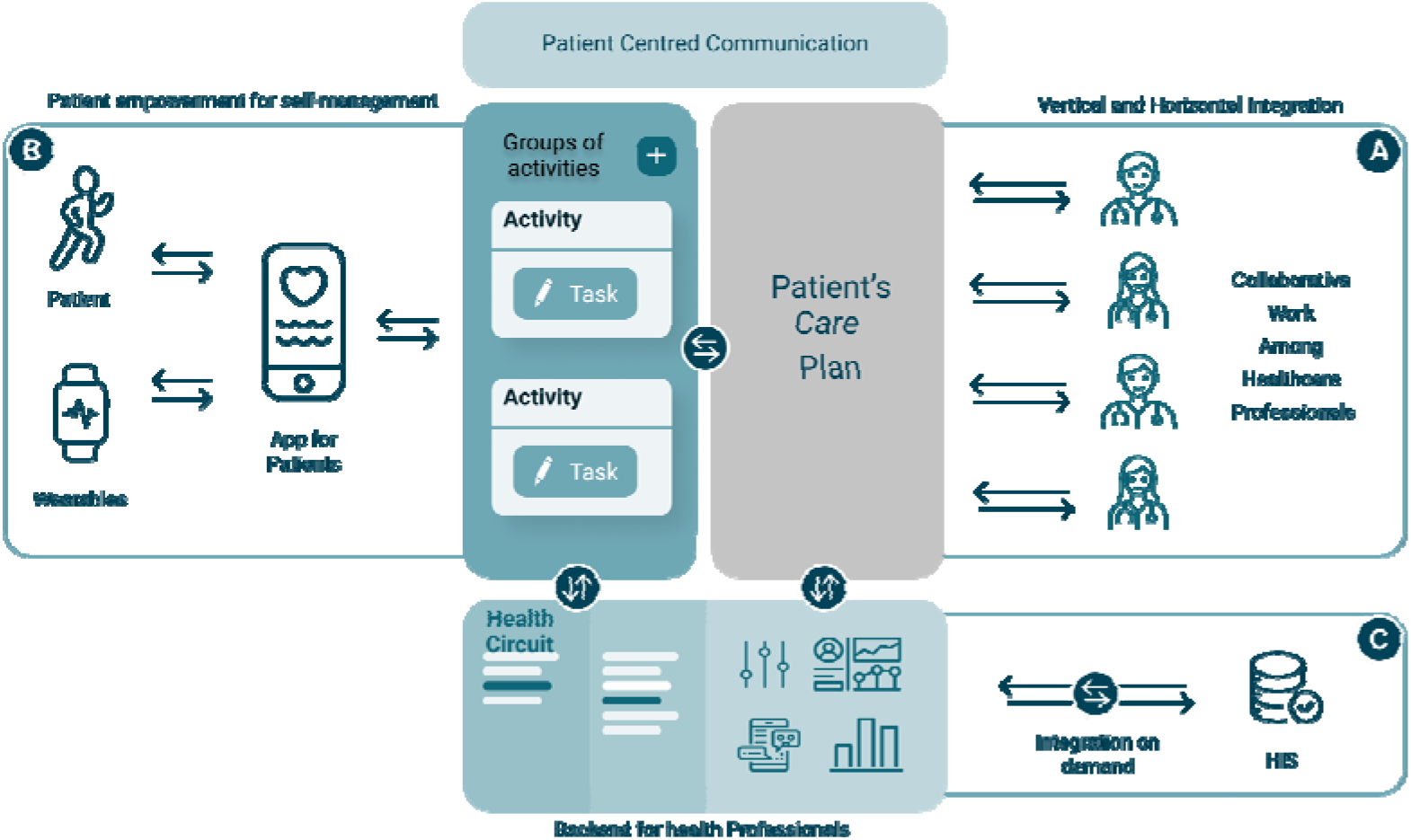
Schematic depiction of Health Circuit ACM approach. **Panel A:** healthcare professionals easily adapt and customize shared care plans over time, facilitating a connected experience for both the patient and the healthcare professionals. **Panel B:** patients participate in effective health coaching and self-management strategies, on any device, for a goal-oriented and personalized health plan to manage both expected and unexpected events. **Panel C:** without requiring tight system integration, Health Circuit allows healthcare providers to progressively replace conventional disease-oriented approaches with a predictive, preventive, personalized and participatory integrated care approach.

-Event driven architecture. It is designed with an Event Driven Architecture in mind. This means that the internal structure of the care plan is based in events and actions (publish-subscribe software architecture pattern) that can be linked together.

- Events. Anything that happens, from “the activity A started” or “the group of activities X ended” to “the weekly total step count passed the 80% threshold”.
- Actions. A limited set of operations that can be performed, like “start/end an activity/activity group” or “send a push/SMS/email notification to a patient/professional”. More advanced scenarios will introduce actions that represents integrations with 3rd party systems like “upload a report to the HIS”.

Such an event driven architecture is scalable and open, and allows introducing a visual workflow editor, such as healthcare professionals without technical skills and define and/or customize their own care plan templates.

-Tailored integration with existing corporate-specific health information systems. Delivered as a low-barrier corporate communication channel with ACM features, without requiring tight system integration, will maximize adoption across the global healthcare IT market. When required, Health Circuit can be easily integrated with site-specific health information systems by means of a standard-based (e.g., HL7-FHIR) interoperability middleware and/or provider-specific web-services/APIs. This can be the case for the calendar of appointments, the agenda of healthcare professionals, admission/discharge reports and/or authentication with corporate-specific centralised user management systems.

### 2.3 Community-based management of complex chronic patients with high risk for hospitalization

The protocol was designed as a cluster randomized controlled trial, conducted by 3 primary care teams, with an intervention to control ratio of 2:1 and follow-up for a period of three months. Eligible patients (n=100) were selected from reference primary care teams within the CAPSBE primary care area of Barcelona.

The inclusion criteria were: (i) Adjusted Morbidity Groups (AMG) score ≥ 3 ^18,19^ as a proxy of moderate to high risk from a multimorbidity criteria; and, (ii) In the intervention group, having a smartphone or tablet compatible with the Health Circuit prototype mobile application and with Internet connection. AMG is an aggregative index which estimates the individual’s burden of disease from a disease-specific weighting obtained from a population-based statistical analysis based on mortality and the use of health services. The exclusion criteria were physical or psychological problems that unable the use of Health Circuit and not having a career.

For both the intervention and the control group, sociodemographic data and digital literacy baseline characteristics were collected at baseline, and the FANTASTIC Questionnaire ^20^ and Elders Health Empowerment Scale (EHES) ^21^ were collected at baseline and at three months, and the patients’ and professionals’ satisfaction and usability of the of the Health Circuit prototype assessed with the self-administration of the Net Promoter Score (NPS) ^22^ and System Usability Scale (SUS) ^23^, respectively. See **Multimedia Appendix** 1 for details.

For the intervention group, a motivational interview was conducted at baseline with one case manager aiming to co-design a personalized and comprehensive care plan, as well as to empower the patient for self-management of his/her conditions (**Table 1S**). The nurse case manager carried out follow-up visits at 15 days, one month and two months after the start of the study and were always available when the patients reported any query or health problem through the app. Patients and professionals pertaining to the intervention primary care centres were instructed on the functionalities of the Health Circuit prototype (**Multimedia Appendix 2**). In the control group, both patients and professionals followed standard of care procedures.

Results are presented as mean (SD) or (%) when indicated. Comparisons were made using chi-square or Fisher’s exact test for categorical variables and Student’s tests, according to the distribution of the variables, for numerical variables.

Study approval was obtained from the Ethics Committee for Clinical Research of Hospital Clinic de Barcelona (HCB/2018/0805) and trial registration NCT04056663 (clinicaltrials.gov, August 14, 2019). Patients read, understood and accepted the informed consent which was signed before enrolment to the study.

### 2.4 Prehabilitation of high-risk candidates for major surgical procedures

A descriptive observational study was carried out including all patients (n=104) who completed the Prehabilitation program at HCB ^8^ between July 2020 and July 2021.

The inclusion criteria were: (i) Having a smartphone or tablet compatible with the prototype mobile application (PREHAB) and with Internet connection. The exclusion criteria were: (i) Physical or psychological problems that unable the use of the mobile application; and, (ii) Not having a career.

The prototype version of the Health Circuit tailored to Prehabilitation process (PREHAB – TRL 7) allowed care team members to prescribe and monitor non-pharmacological interventions for patients’ self-management through a mobile application, including: i) Physical activity goals; i) Nutritional tips; iii) Mindfulness exercises; and iv) Patient reported outcomes. Moreover, patients had access to a one-to-one chat with the case manager^24^ for enhanced management of multimorbidity. See **Multimedia Appendix 3** for details.

At the end of the Prehabilitation program (4-6 weeks in average), the patient’s satisfaction and usability of the mobile application was assessed with the self-administration of the NPS and SUS, respectively. Moreover, the patient’s usage of the mobile application was evaluated according to: i) The number of chat interactions, ii) Adherence to reporting of daily physical activity, iii) Completion of follow-up questionnaires and iv) Access to the tailored education material on mindfulness, nutrition and physical activity. Results are presented as mean (SD) or (%) when indicated.

In this study the backend for healthcare professionals was not assessed because the recruitment and follow-up of study participants was done with only one healthcare professional, a case manager physiotherapist.

The Ethics Committee for Medical Research of HCB approved the study (HCB/2016/0883). The informed consent was understood, accepted, and signed by all subjects included.

## 3. Results

### 3.1. Community-based management of complex chronic patients with high risk for hospitalization

The study was conducted between September 2019 and March 2020. Two primary care units were randomly assigned as intervention and 1 as a control unit. From an initial sample of 100 eligible patients, 41 did not meet the participation criteria (see CONSORT study flowchart in Multimedia Appendix 4). The remaining 59 patients were assigned to one of the study arms according to the reference center. Up to 47 patients, 31 in the intervention arm and 16 controls, completed follow-up. The initial demographic characteristics are shown in Table 1.

**Table 1.** Sociodemographic baseline characteristics of 47 patients with high risk for hospitalization

|  | **Intervention (n=31)** | **Control (n=16)** | **P<0.05** |
| --- | --- | --- | --- |
| Age, mean (SD) | 72.7 (13.7) | 78.1 (8.8) |  |
| Male, gender, n (%) | 20 (64.5) | 12 (75.0) |  |
| Educational superior level, n (%) | 19 (61.3) | 6 (37.5) |  |
| Widowed Status, n (%) | 14 (45.2) | 1 (5.9) | p<0.004 |
| Living with family, n (%) | 22 (71.0) | 14 (87.5) |  |
| Retired status, n (%) | 20 (64.5) | 15 (94.1) |  |
| AMG<sup>1</sup> score, n (%) |  |  |  |
| Moderate risk [$P_{80} - P_{95}$] | 15 (48.4) | 7 (43.8) | |
| High risk [$P_{95} - P_{99}$] | 16 (51.6) | 9 (56.2) | |

1. AMG, Adjusted Morbidity Groups, index indicating multimorbidity and complexity of the patients<sup>25,26</sup>

Detailed information on availability of smartphones (intervention 88% and controls 77%), digital baseline characteristics, baseline use of technologies and health information sources can be found in Multimedia Appendix 5.

During the study period, a total of 28 consultations, 12 clinical and 16 administrative, were done by 26 out of 31 patients of the intervention group. In all cases, the events were timely solved. The offline chat was used in 78% of the consultations and the phone in the remaining 22%.

The intervention had positive clinical impacts in terms of less use of healthcare resources (NS), increase in patient empowerment (p<0.001), and improvement in perception of continuity of care (p<0.001). As depicted in Table 2, the intervention group had fewer hospital and primary care emergency room visits, lower hospitalization rate, less mortality and half of visits to primary care than the control group. However, the results were not statistically significant.

**Table 2.** Clinical and patient-reported outcomes during study the study period.

| **Outcome variables** | **Intervention**<br>**(n=31)** | **Control**<br>**(n=16)** | **P<0.001** |
| --- | --- | --- | --- |
| **Hospital emergency room visits, n** | 0 | 1 |  |
| **Primary care emergency room visits, n(%)** | 4(13) | 7(44) |  |
| **Primary care visits, n (mean per patient)** | 51 (1.59) | 52 (3.25) |  |
| **Hospitalizations, n(%)** | 0(0) | 2(13) |  |
| **Mortality, n(%)** | 0(0) | 2(13) |  |
| **Baseline live style<sup>20</sup>, m(SD)** | 41.18(3.53) | 36.06(4.36) |  |
| **Δ live style (pre-post)<sup>20</sup>, m(SD)** | 2.62 (4.49) | 0.43(4.15) |  |
| **Baseline empowerment<sup>27</sup> m(SD)** | 36.46(5.38) | 28.44(6.24) | p<0.001 |
| **Δ empowerment (pre-post)<sup>27</sup>, m(SD)** | 4.5(5.23) | -4.56(5.38) | p<0.001 |
| **Continuity of Care<sup>28</sup>, m(SD)** | 4.84(2.40) | 2.75(1.26) | p<0,001 |

#### 3.1.1. Usability and acceptability

The patients reported good satisfaction and usability of the mobile application (NPS score 31 and SUS 68/100), as well as a high acceptance rate (mean score of 7.8/10). However, the backend for health professionals scored neutral in terms of satisfaction and usability (NPS score -80 and SUS 54/100) with an acceptance score averaging 5/10). Despite such results in terms of acceptability/usability of the backend, 75% of the professionals participating in the study indicated a good overall quality of the remote consultations, 86% acknowledged that they will continue to use remote consultations as a support tool and 71% of the professionals considered that telemedicine can improve the state of patients’ health.

### 3.2. Prehabilitation of high-risk candidates for major surgical procedures

A total of 104 patients (mean age 69 ±11 years, 67% men) agreeing to download the mobile application (PREHAB - TRL 7)^29,30^ during the study period were included in the analysis of patients’ usability and acceptability of the application.

The patient’s mobile application obtained a good usability score (NPS score 40 and SUS 85/100), as well as a high acceptance rate (mean score of 8.4/10).

The patient’s primary use of the mobile application was to follow the prescription of daily physical activity, in 96% (n:100) of the cases. The weekly nutrition follow-up questionnaires were completed by 40% (n:42) of the patients. Up to 37% (n:38) of study participants used the chat at least once to communicate with the prehabilitation team members (i.e., physiotherapists, nutritionists, etc.). Only 21% (n:22) of the participants completed the daily mindfulness exercises. Educational tips on nutrition and physical activity were consulted by 33% (n:34) and 26% (n:27) of the study participants, respectively.

## 4. Discussion

### 4.1. Principal findings

We have presented the Health Circuit ACM approach to help healthcare professionals create patient-centered workflows combining both pharmacological and non-pharmacological interventions. It functions as a semi-automated task manager for professionals and patients, allowing them to perform certain activities as part of the patient’s action plan. The approach encourages collaboration and sharing of knowledge among healthcare professionals. The feasibility of the approach have been assessed in two innovative healthcare services for: (i) Enhanced management of complex chronic patients with high risk of hospitalization through a secure bi-directional communication channel; and, (ii) Scalability of a prehabilitation service, that have previously demonstrated to be efficient ^7^ and cost-effective ^8,17^, incorporating the virtual sharing of health plans that include non-pharmacological activities toward patient empowerment for self-management.

The pilot study on community-based management of complex chronic patients with high risk for hospitalization (TRL 5) showed the potential of the approach to enhance management of unplanned health events, leading to less use of healthcare resources, as well as to improve patient empowerment for self-management and perception of continuity of care. The results are encouraging. Specially, when considering that both patients and healthcare professionals reported a reasonably good usability and acceptability, even though the high mean age of study participants (72.7 years) and the series of limitations that prototype-level digital health tools of such low TRL tend to have. Similarly, good usability and acceptability of the ACM approach (TRL 7) were reported in the pilot study on prehabilitation of high-risk candidates for major surgical procedures, which resulted in patients’ adherence to the non-pharmacological treatment, especially in the case of daily physical activity.

### 4.1. Strengths and weaknesses

Besides the pre-commercial nature of the digital health tools supporting each one of the two feasibility studies, reasonably good usability and acceptability results were obtained in real world clinical settings. Nevertheless, the positive results reinforced the need to pursue the development of a commercial product with potential for cost-effectiveness and profitability in re-al-world settings.

Although one of the limiting factors for adoption of value-based care services is still stakeholders’ acceptability, and usability - especially for the elderly - the current study contributed to the growing body of evidence that indicate positive effects of eHealth services on patient health outcomes ^31–34^ (e.g., by enhanced patient empowerment and/or adherence to care plans) at a lower cost.

### 4.2. The Health Circuit approach to ACM

Change management is critical for ensuring effective digital health transformation, as it helps stakeholders adapt to new processes and technologies, build support for change, and achieve the goals of the transformation. By taking the ACM approach of Health Circuit, healthcare organizations can minimize disruption, ensure stakeholder buy-in, and improve the quality of care. Briefly, the front-line support to flexible case-centered collaboration is a secure multichannel communication, whereas shared care plans can be introduced to multidisciplinary care teams in a stepwise manner leading to a more standardized care protocol as the result of an effective adoption of organizational interoperability.

Therefore, the Health Circuit approach to ACM can play a crucial role for the adoption of integrated care by enabling healthcare professionals from different disciplines and organizations to collaborate and share knowledge, which can lead to better-informed decisions about the patient care. Hence, also playing a significant role in supporting personalized medicine by enabling healthcare professionals to create and adapt individualized treatment plans to the specific needs and circumstances of each patient.

### 4.3. Perspectives and future developments

The aim of the Health Circuit approach to ACM is to facilitate large-scale adoption of value-based care services, such us the two innovative healthcare services considered in the current study. Therefore, future perspectives focus in easing the implementation of digi-tally enabled integrated person-centred care by means of a flexible approach to shared care processes and a holistic approach to health risk assessment.

In this vision, ACM processes dictate information needs, which is a great opportunity to decouple data from applications, enabling data to follow patient-centric care processes and helping to scale-up innovations for value-based healthcare. Storing data with open standards, such as OpenEHR, data analysts can make a secondary use to understand cost-effectiveness within the context of a certain care plan. With this infostructure, AI ex-perts can train automated decision support for health risk assessment. This can potentially be also done for health risk assessment, to indicate what can be the best care plan fit for a given type of patient - the one that minimizes certain risks, like mortality or unplanned hospital admissions.

## 5. Conclusions

The Health Circuit approach to ACM demonstrated potential for value generation, as well as reasonably good usability and acceptability results by end users. The two pilot studies allowed to validate the feasibility of the approach to enable collaboration and knowledge sharing among healthcare professionals from different levels of care and with patients, which can lead to better-informed decisions about the patient care.

Therefore, the Health Circuit approach to ACM can play a crucial role for the adoption of integrated care in a flexible and tailored way, depending on the digital health transformation maturity of the end users.

## Data Availability

All data produced in the present study are available upon reasonable request to the authors

## Supplementary Materials

The following supporting information can be downloaded at: www.mdpi.com/xxx/s1, Figure S1: Flow Diagram Health Circuit, Figure S2: Baseline us of technologies; Table S1: Digital baseline characteristics in the intervention group.

## Author Contributions

Conceptualization, C.I. and R.J.; Methodology, H.C. and C.I.; Software, C.I.; Validation, H.C., C.I., M.L., D.F., S.A. and R.J.; Formal Analysis, C.I. and H.C.; Investigation, D.F, M.L. and H.C.; Resources, C.I. and R.J.; Data Curation, H.C., C.I., M.L., D.F., S.A. and R.J.; Writing – Original Draft Preparation, H.C., C.I. and R.J.; Writing – Review & Editing, H.C., C.I., M.L., D.F., S.A. and R.J.; Visualization, H.C., C.I., M.L., D.F., S.A. and R.J.; Supervision, R.J.; Project Administration, C.I.; Funding Acquisition, R.J.

## Funding

The JADECARE project is a research and innovation project funded by the European Union’ Horizon 2020 Research and Innovation Program under grant agreement number 951442. The information reflects only the authors’ views, and the European Commission is not responsible for any use that may be made of the information.

## Institutional Review Board Statement

The study was conducted according to the guidelines of the Declaration of Helsinki and approved by the Institutional Review Board (or Ethics Committee) of Hospital Clinic (protocol code HCB/2018/0805 and HCB/2016/0883) and date of approval.

## Informed Consent Statement

Informed consent was obtained from all subjects involved in the study.

## Conflicts of Interest

I.C. and J.R. has shares of Health Circuit SL, a spin-off company of Hospital Clinic of Barcelona. All other authors declare no conflict of interest.

## Notes

### Competing Interest Statement

The authors have declared no competing interest.

### Clinical Trial

NCT04056663

### Author Declarations

The Ethics Committee for Medical Research of HCB approved the study (HCB/2016/0883 and HCB/2018/0805). The informed consent was understood, accepted, and signed by all subjects included.

