## Supplementary Materials for "Health Circuit: a practice-proven adaptive case management approach for innovative healthcare services"

**APPENDIX 1**

**NPS and SUS measures**

The Net Promoter Score is a known questionnaire used to assess satisfaction with a product, which includes a key question: “How likely is it that you would recommend our system to a family member or friend?”. Patients can give an answer ranging from 0 (“not at all likely”) to 10 (“extremely likely”). Individuals scoring a 9 or a 10 are called “promoters”, individuals scoring 7 or 8 are called “passives” (or neutrals) and individuals scoring 0 to 6 are labelled as “detractors”. The SUS was developed by John Brooke in 1986 and consists of a 10-item questionnaire scored on a 5-point Likert scale from 0 (strongly disagree) to 5 (strongly agree). The overall score is calculated from a sum of all item scores multiplied by 2.5 and can range from 0 to 100. A system or product that received score of 68 and above is considered to have good usability

**APPENDIX 2**

**User manual: Community-based management of complex chronic patients with high risk for hospitalization**

**• Language selection:** As a user, you can select between three languages: Spanish, Catalan and English.

**• Tutorial :** As a user, when I download the app , I can see a short tutorial of the main functionalities that the app offers .

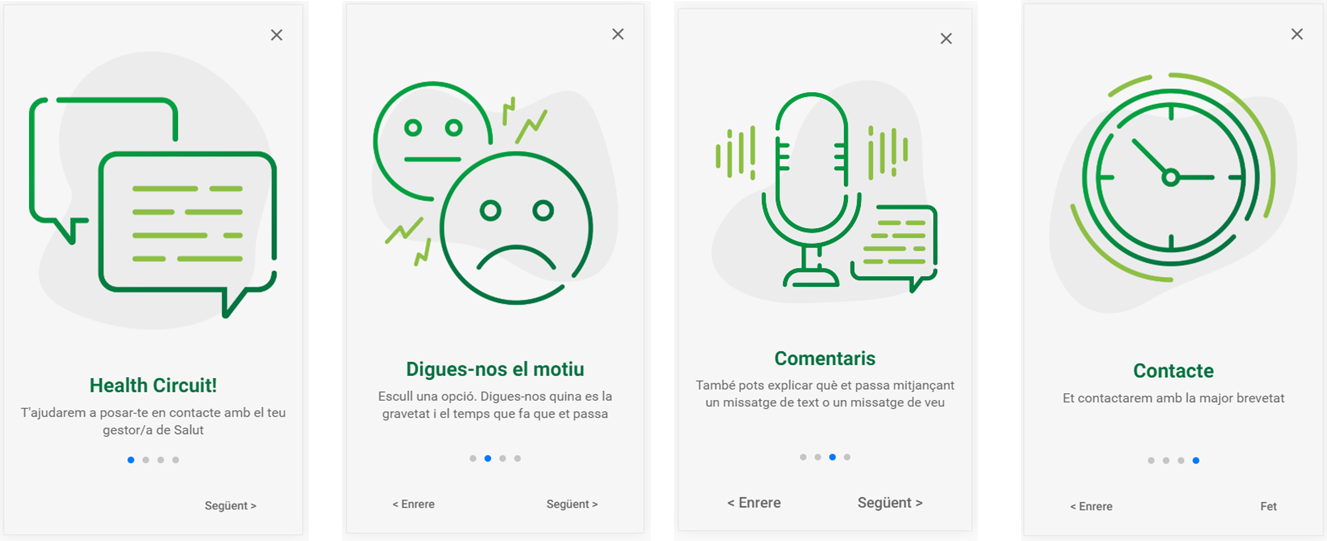

- **Login:** As a user I must access with an email and password provided by the clinic. In the HealthCircuit app, I can view the key to reduce the risk of entering an incorrect password.
  - Particularities: You are only asked to enter the username and password the first time you access 🡪The user must be aware that when logging in he/she consents to the data of the connection being saved (through consent)
    - As long as the user does not log out, he will be able to access Health Circuit without the need to enter a password.
    - If the user logs out, he must re-enter the username and password. In addition, while you are not connected to Health Circuit, you will not be able to receive communications from the clinic (messages or calls), you will only be able to see the messages received once you log back in as a user.
    - The user must be informed about:
      - The Hospital Clínic has a database with logins and passwords pseudonymized _
      - You only need to enter the login the first time you access the app. Therefore, in case of loss, theft, etc., it is recommended to notify the Clinic immediately.
      - Phase I: All the information sent is registered in Circuit. The user must be informed of the storage time of the conversations. To comply with GDPR, any data we save (conversations in circuit) must be based on the legitimacy of the treatment (Why we save it and the knowledge of the interested party about the treatment and retention of data).
      - Phase II: By incorporating the chat option, all the information sent through triage is recorded both in Circuit and on the user's device. To comply with GDPR, any data we save (conversations in circuit) must be based on the legitimacy of the treatment (Why we save it and the knowledge of the interested party about the treatment and retention of data).
    - If the user has forgotten the password, they can request a reminder by calling the Clinic.

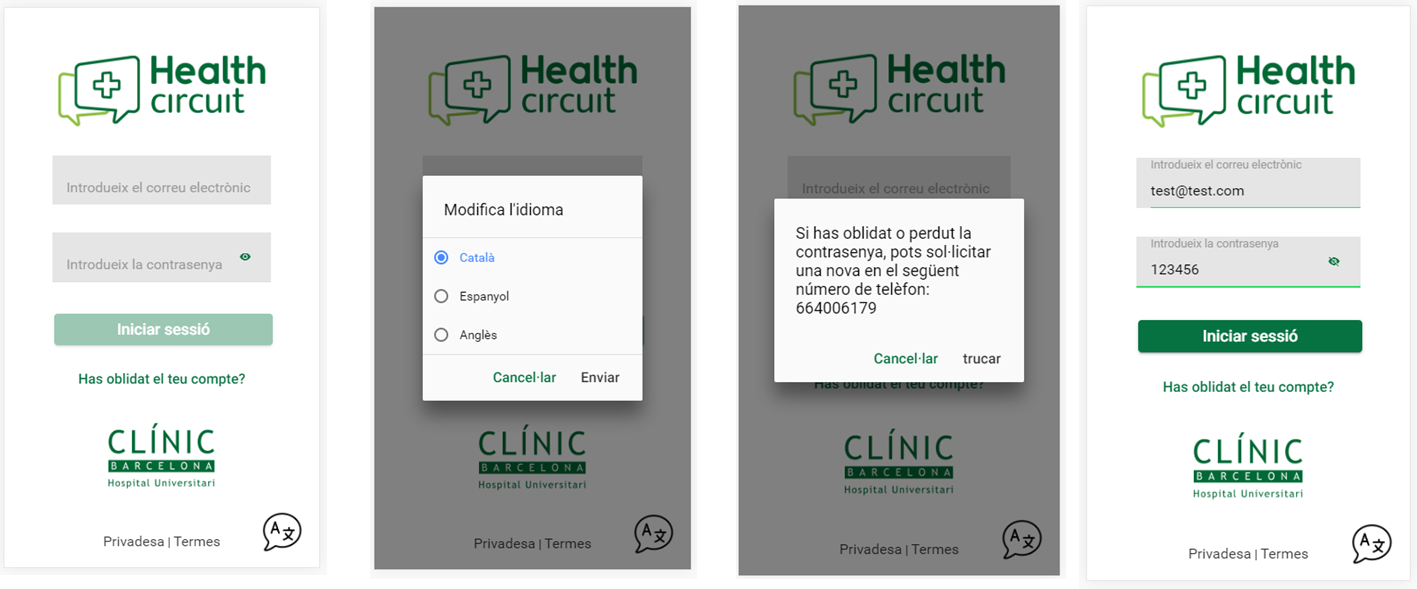

- **Triage:**
  - As a user, I can select from the following options: Type of inquiry: Pain, Shortness of breath, Fever, Other complaints or Administrative doubts *.
  - Symptom intensity **: Very intense, intense, neutral, mild or very mild.
  - Time since symptom onset **: Less than 1h, between 1h-12h, between 12h-24h, between 2-7 days, More than 1 week, More than 1 month.
  - Optional: The user is given the possibility to detail the reason for the query by recording an audio or sending a text message.

* For the "Administrative Doubts" option, the intensity or time since symptom onset screens are not generated. The user is redirected directly to the audio or text registration screen.

** Both sections are mandatory to enter to go to the next screen.

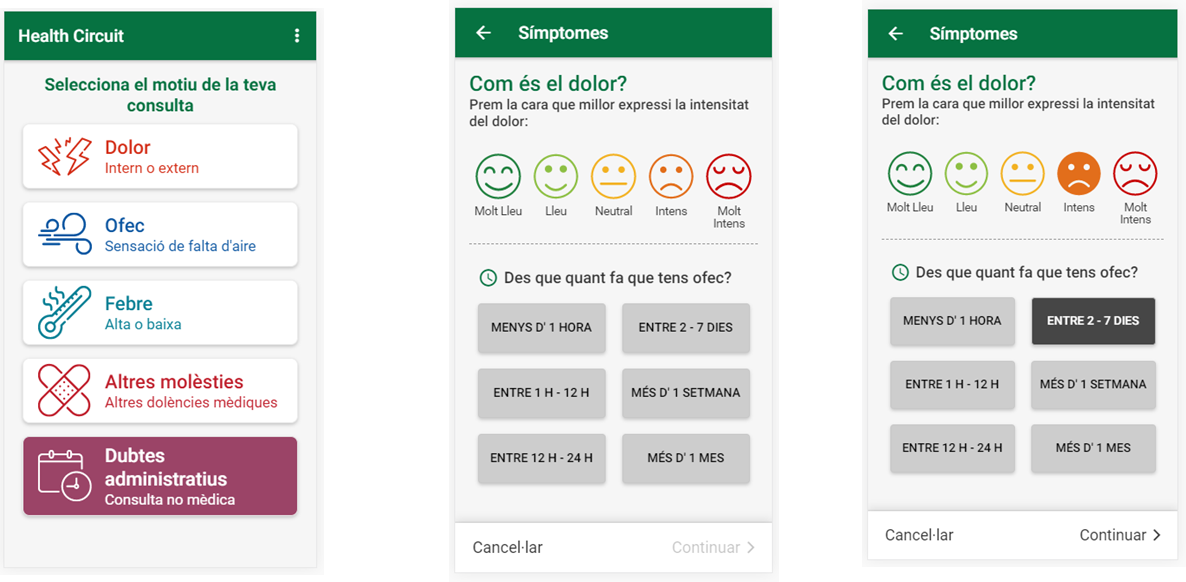

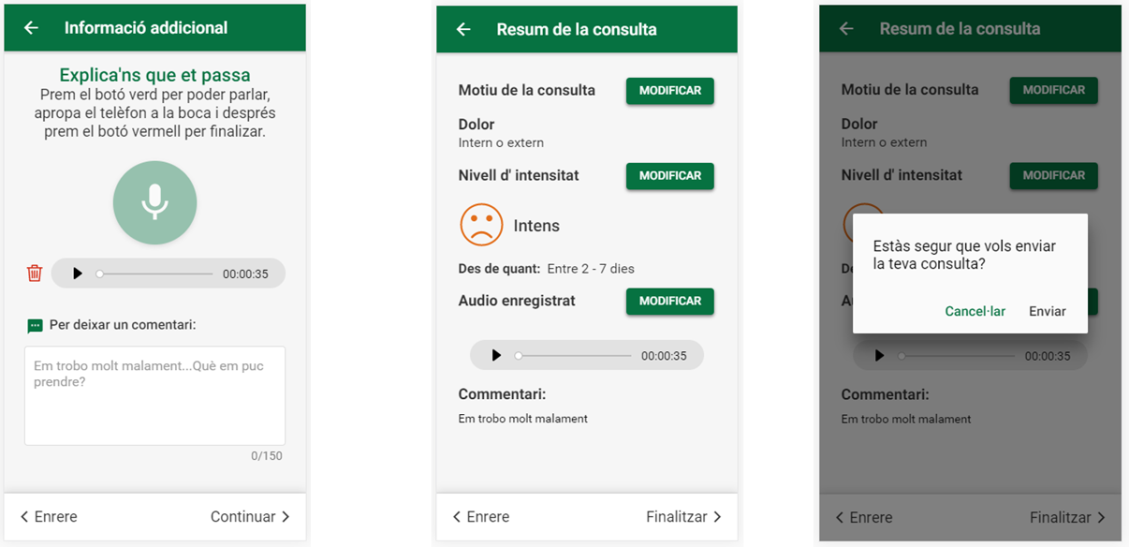

- **Chat:**
  - Once the triage has been completed and sent, the "Manager / a not available" screen will automatically appear: Your health manager / a will contact you within 3 hours at the most (from Monday to Friday from 08:00 a.m. to 08:00 p.m.). If you cannot wait, we recommend that you call 061 or go directly to your usual health center. Additionally, you can contact us again via chat enabled for any query related to the sent triage status. "

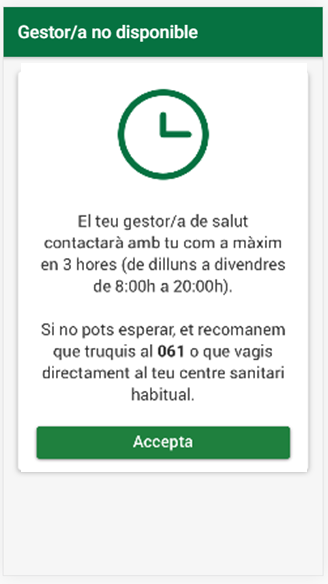

- As a user, upon acceptance, I will automatically be redirected to the initial triage screen where I will have chat enabled. I will be able to access the chat icon and type any additional details about the submitted triage (Example: My condition has worsened). I will also be able to receive triage-related messages from the Clinic when the health manager is available, within a maximum of 3 hours from the sending of the triage by the user.
- If I wish to create a new triage again, a message will appear when selecting any of the 5 options, stating " Are you sure you want to create a new triage? Creating a new triage means that the chat conversation associated with the last triage will be automatically removed. " You can select "Yes" or "Back"
- If I select "Yes", the chat from the last triage will be deleted and a new one will be created when the triage submission is complete.
- The Clinic, for its part, through Circuit, will not be able to reply to triages sent previously, it will only be able to reply about the last triage.
- **Push notifications:**
  - As a user, as long as I have not logged out of the application, I may receive notifications of incoming calls / video calls or incoming messages, even if the application is not open in the foreground.
  - If the user has logged out, notifications will not be received. You will only be able to view the messages received through the chat when you log in again.
    - Particularities:
      - The user, through the signed consent, must authorize receiving calls / messages in Push notifications. A settings section where you can activate / deactivate notifications is not included. Therefore, you must be aware that if you do not wish to receive push notifications, you must contact the Clinic.
- **The Clinic may contact the participant regardless of whether there is an event or not.**
  - At any time and regardless of whether or not a triage has been created, the Clinic can contact the user.
  - For this and with the aim of complying with GDPR, it is necessary for the user to consent and to know for what reason they may be contacted. You must include in the terms of privacy and in the consent to be signed by the user for which purpose he can be contacted and accept it.

**APPENDIX 3**

**User manual: Prehabilitation of high-risk candidates for major surgical procedures**

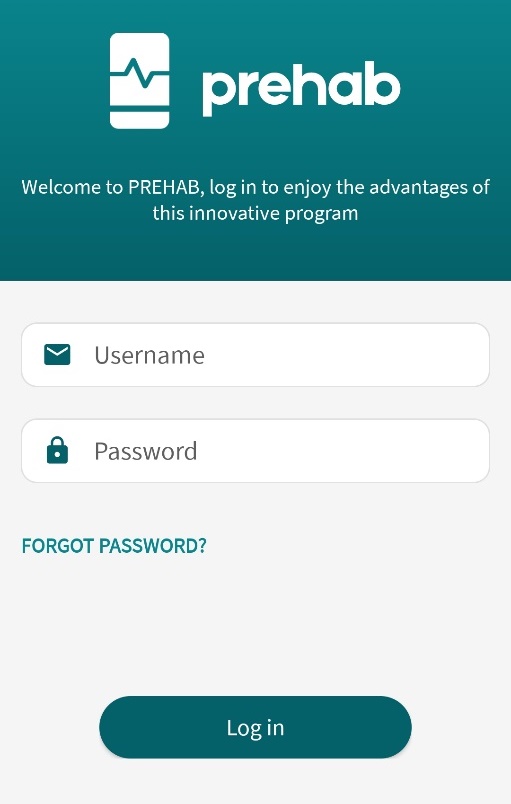

**Login**

To use Prehab, the app user’s credentials must be validated by following the instructions from the clinician.

You will need to enter your **username** and **password** for access.

On the same screen you’ll see the **“Forgot password?”** option, so it can be changed if you've forgotten it.

In addition, you can view the app’s **terms and conditions**.

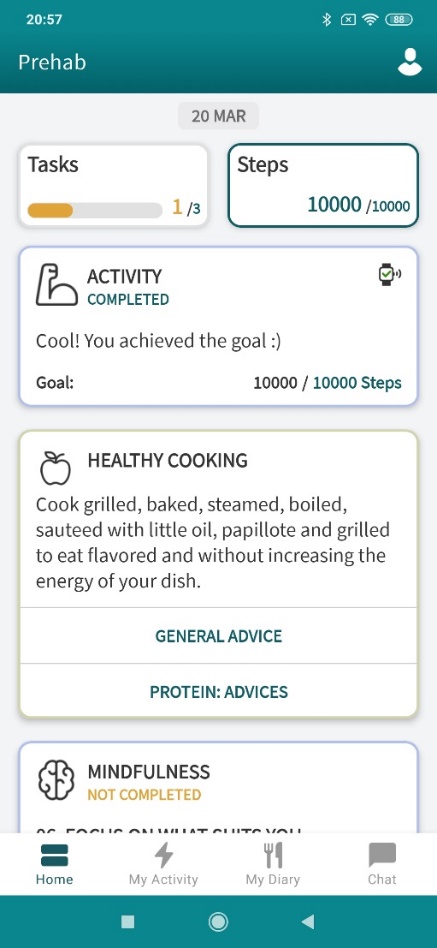
**Home screen**

This is the screen you’ll see when you start a session as a patient. It contains the following information:

The **Profile** icon is at the top; it can be used to view your user information or information about the connection with your activity wristband, and it also allows you to log out.

Beneath it, you’ll see two fields: the **Tasks** field shows the number of activities to be completed during the day, and the **Steps** field shows the number of steps to be taken.

The other fields refer to instructions received from clinical specialists. There are also fields with resources for the Prehab programme and a welcome video.

The lower menu shows the following options: **Home**, to return to the home screen; **My Activity**, where you can more closely monitor your physical activity; **My Diary**, where you can take and display photos of what you've eaten each day; and **Chat**, where you can start a chat with clinicians.

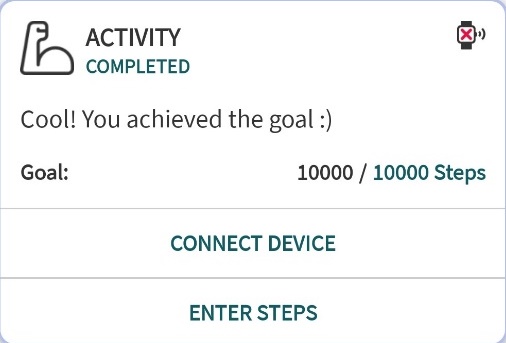
**Activities**

The following activities may be visible for the patient:

**Physical activity:** This lets you know how many steps to take, and gives you the option to enter steps manually if you don’t have an activity wristband. You can also connect your device to measure your steps automatically.

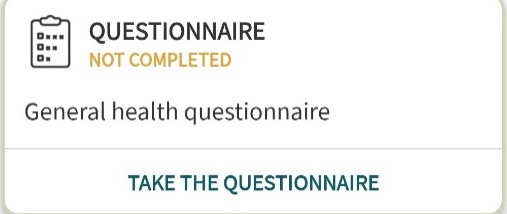

**Questionnaires:** The type of prescribed questionnaire is displayed, and you have the option to complete it. Once you've entered your answers, the status will change from “Not completed” to “Completed,” and you can view your responses.

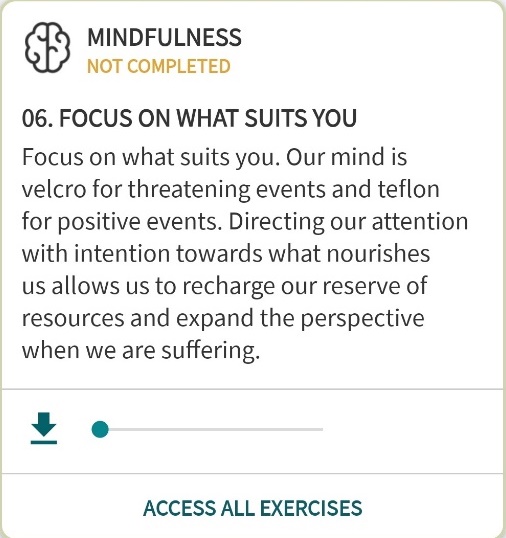

**Mindfulness:** An introductory text is displayed, and you’re given the option of completing the day’s proposed mindfulness exercise. You also have the option of viewing all the exercises in case you want to do more.

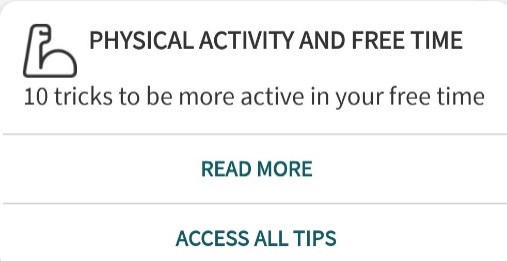

**Physical activity advice:** Each day a different piece of advice from the clinicians will be displayed. You’ll also be able to view all the advice if you want more information.

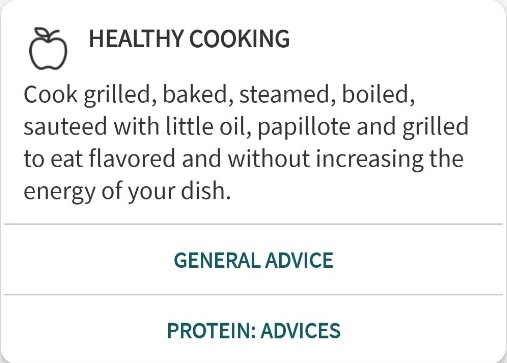

**Nutrition advice:** This section contains nutrition advice provided by the clinicians and will be a great help to patients. A different piece of advice will be posted each day.

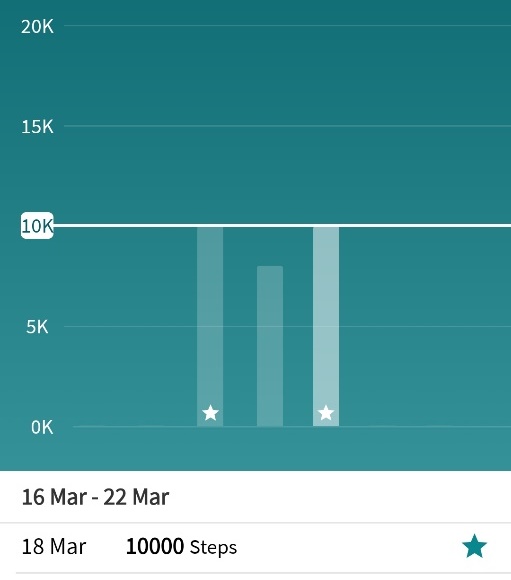

**My activity**

On this screen you can monitor your physical activity more closely with the aid of a bar graph showing the number of steps taken each day.

A white horizontal line shows your goal so you can easily see whether you've reached it.

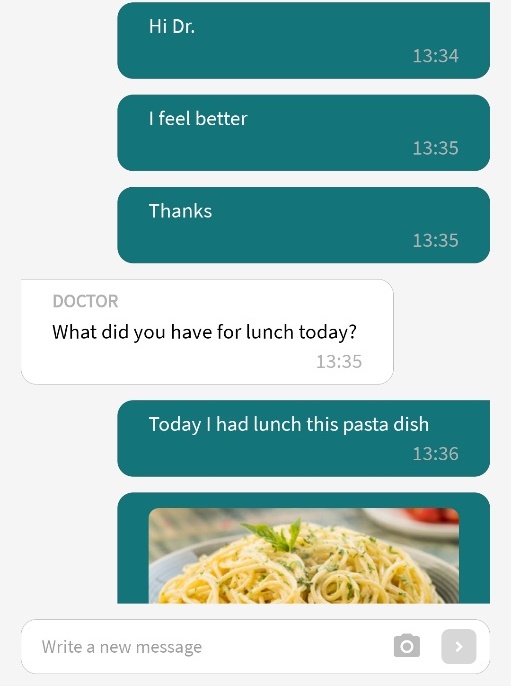

**Chat**

On the Chat screen you can quickly and easily exchange messages with clinicians. You can also send clinicians a photo stored on your mobile or a photo taken with the camera.

Clinicians can also send PDF documents, which will appear on the chat screen so the patient can read them.

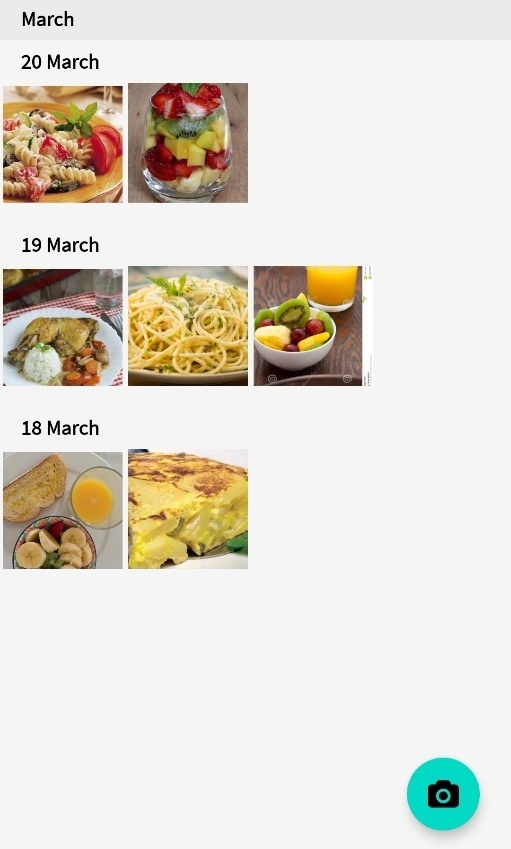

**Food diary**

This option appears only if the clinician has prescribed a food diary for the patient.

This screen allows patients to send photos of what they've eaten during the day so that the clinician can monitor their diet.

The various photos taken each day can also be viewed, with the most recent ones appearing first.

**APPENDIX 4**

**Figure S1. Flow Diagram Health Circuit**

**CONSORT 2010 Flow Diagram Health Circuit**

Allocated to control group (n=18)

- Received allocated intervention (n=18)
- Did not received allocated intervention (n= 0)

Allocated to intervention group (n=41)

- Received allocated intervention (n=39)
- Did not received allocated intervention (n= 2)
  - Technological problems (n = 2)

### Allocation

Excluded (n=41)

- Died (n= 5)
- Did not answer telephone (n=5)
- Not meeting inclusion criteria(n=20)
- Refuse (n= 11)

Center randomized

Lost to follow-up (n=2)

- Died (n=2)

Total (n=16)
♦ Excluded from analysis (n=0)

### Follow-Up

Lost to follow-up (give reasons) (n=8)

• Technological problems (n = 2)

• Died (n = 1)

• Worsening of health (n = 2)

• Lost mobile device (n = 1)

• Unreachable (n = 2)

### Analysis

Total (n=31)
♦ Excluded from analysis (n=0)

### Enrollment

Assessed for eligibility (n=100)

**APPENDIX 5**

**Table S1**. Digital baseline characteristics in the intervention group.

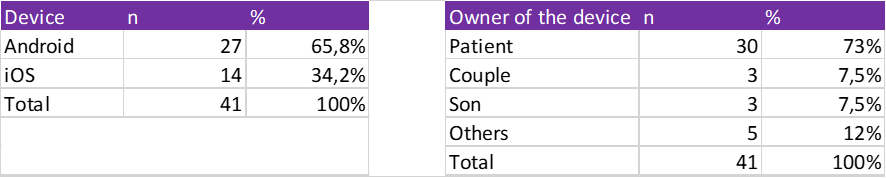

**Figure S2.** Baseline use of technologies and health information sources

Health information sources

|  | Intervention group | Control group |
| --- | --- | --- |
| Healthcare professional | 100% | 100% |
| Pharmacist | 58.0645% | 62.5% |
| Friends and family | 45.1613% | 31.25% |
| Mass media | 22.5806% | 25% |
| Internet | 51.6129% | 18.75% |
| Social media | 0% | 0% |
| Apps | 9.67742% | 0% |

Devices that use (%)

|  | Intervention group | Control group |
| --- | --- | --- |
| Computer | 70.7317% | 50% |
| Smartphone with internet | 87.8049% | 72.2222% |
| Tablet | 51.2195% | 44.4444% |
| Devices for health (smar watch, wristband activity…) | 12.1951% | 11.1111% |
